# Sentiment Informed Timeseries Analyzing AI (SITALA) to curb the spread of COVID-19 in Houston

**DOI:** 10.1101/2020.07.22.20159863

**Authors:** Prathamesh S. Desai

## Abstract

Coronavirus disease (COVID-19) has evolved into a pandemic with many unknowns. Houston, located in the Harris County of Texas, is becoming the next hotspot of this pandemic. With a severe decline in international and inter-state travel, a model at the county level, as opposed to the state or country level, is needed. Existing approaches have a few drawbacks. Firstly, the data used is the number of COVID-19 positive cases instead of positivity. The former is a function of the number of tests carried out while the latter is normalized by the number of tests. Positivity gives a better picture of the spread of this pandemic as with time more tests are being administered. Positivity under 5% has been desired for the reopening of businesses to almost 100% capacity. Secondly, the data used by models like SEIRD lacks information about the sentiment of people with respect to coronavirus. Thirdly, models that make use of social media posts might have too much noise. News sentiment, on the other hand, can capture long term effects of hidden variables like public policy, opinions of local doctors, and disobedience of state-wide mandates. The present study introduces a new AI model, viz., Sentiment Informed Timeseries Analyzing AI (SITALA), that has been trained on COVID-19 test positivity data and news sentiment from over 2750 news articles for the Harris county. The news sentiment was obtained using IBM Watson Discovery News. SITALA is inspired by Google-Wavenet architecture and makes use of TensorFlow. The mean absolute error for the training dataset of 66 consecutive days is 2.76 and that for the test dataset of 22 consecutive days is 9.6. The model forecasts that in order to curb the spread of coronavirus in Houston, a sustained negative news sentiment will be desirable. Public policymakers may use SITALA to set the tone of the local policies and mandates.

**One Sentence Summary:** An AI model based on news sentiment and COVID-19 test positivity is developed to predict the spread of coronavirus in Houston and to guide future local public policy.

## Background, problem, and solution

In the present-day USA, pandemic models of COVID-19 at the level of state or country *(1-4)* are not of much use due to a severe decline in air travel *(5,6)*. The most predominant spread of the virus is then restricted to the geography of a county or a couple of neighboring counties. National, and more so local, news articles provide a fairly accurate picture of the ongoing situation in a crisis-stricken county, Harris in the case of the present study. Local public policy makers can thus play an important role to set the tone or sentiment of the news at the county level *(7).* The spread of coronavirus in the short-term future is a function of test positivity data, however, over longer time periods, news sentiment starts playing an important role. For example, a stay-at-home order, with a strong negative sentiment, issued today may only start seeing the decline in test positivity after 10-14 days due to the incubation period of the coronavirus. Existing approaches have a few drawbacks *(2,8,9,10)*: Firstly, the data used is the number of COVID-19 positive cases instead of positivity. Secondly, the data used by models like SEIRD lacks information about the sentiment of people with respect to coronavirus. Thirdly, models which make use of social media posts might have too much noise. This study attempts to develop a multivariate artificial intelligence (AI) model to analyze timeseries of COVID-19 positivity and news sentiment. The AI model is inspired by Google’s Wavenet *(11)* architecture and uses IBM Watson Discovery News *(12)* to mine COVID-19 sentiment in the news articles.

## Data

The COVID-19 test positivity data for Harris county was obtained from the website of Texas Department of State Health Services (https://dshs.texas.gov/coronavirus/additionaldata.aspx). Couple of instances of bad or missing data were filled using linear interpolation. IBM Watson Discovery was used to mine the news sentiment in **2867 news articles** over the period of 3 months. The query used in this study is provided in the supplementary material. The entire dataset is also provided in the supplementary material.

## Model *(11-16)*

Daily COVID-19 positivity rate and news sentiment are passed to a Wavenet-inspired multivariate CNN to predict the future COVID-19 positivity rate. The AI is named **SITALA** or <u>S</u>entiment <u>I</u>nformed <u>T</u>imeseries <u>A</u>nalyzing <u>A</u>I. A 16-day window, based on coronavirus incubation period of 14 days, with a stride of 1 was used to generate training and test datasets. The architecture of SITALA is shown in **Fig. 1**. Dilated causal convolutions help with the transmission of long-term effects. The output is a single point prediction of COVID-19 in the future. Python code for the neural network architecture is provided in the supplementary material.

**Fig. 1.**
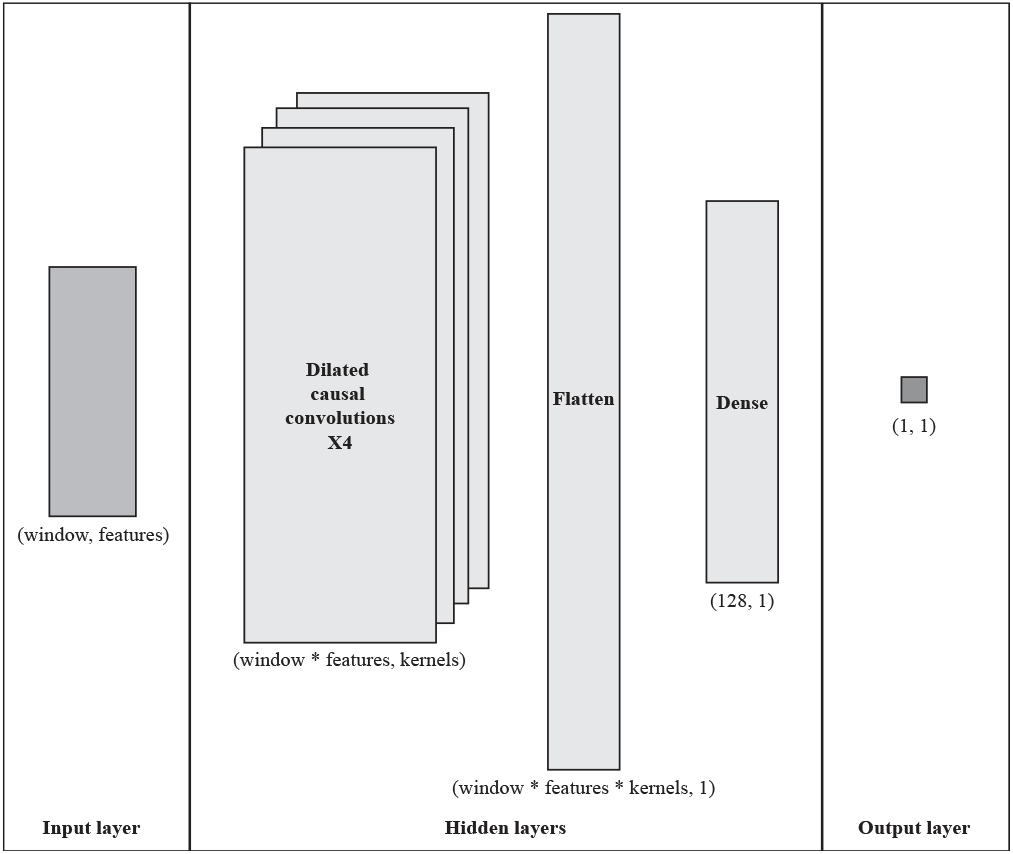
Architecture of SITALA: It is a sequential model that takes a window of multivariate (viz., COVID test positivity and news sentiment from IBM Watson News Discovery) timeseries and outputs the COVID test positivity at the next timestep. This architecture is inspired by Google’s Wavenet *(11)* architecture (viz., dilated causal convolutions). In the present study, a window size of 16 was used and dilations of 1, 2, 4, 8 were used.

## Results

The data used in the present study is shown in **Fig. 2** along with the predictions of SITALA. Data for the Harris county from 04/21 to 07/17 has been used in this study. Number of news articles returned by IBM Watson Discovery News is shown with bars that are multiplied by the sign of the average daily news sentiment. The average daily news sentiment (connected blue squares) can be +1.0 for maximum positive sentiment and -1.0 for maximum negative sentiment. Overall, on most of the days the news sentiment about the spread of coronavirus in Houston, Harris county, Texas has been negative. A big positive spike in news sentiment is seen around the time of social unrest in Houston (05/30 to 06/02). The focus of news might have shifted away from COVID-19 during this timeframe. An upward trend is visible in the COVID-19 positivity data (connected red dots).

**Fig. 2.**
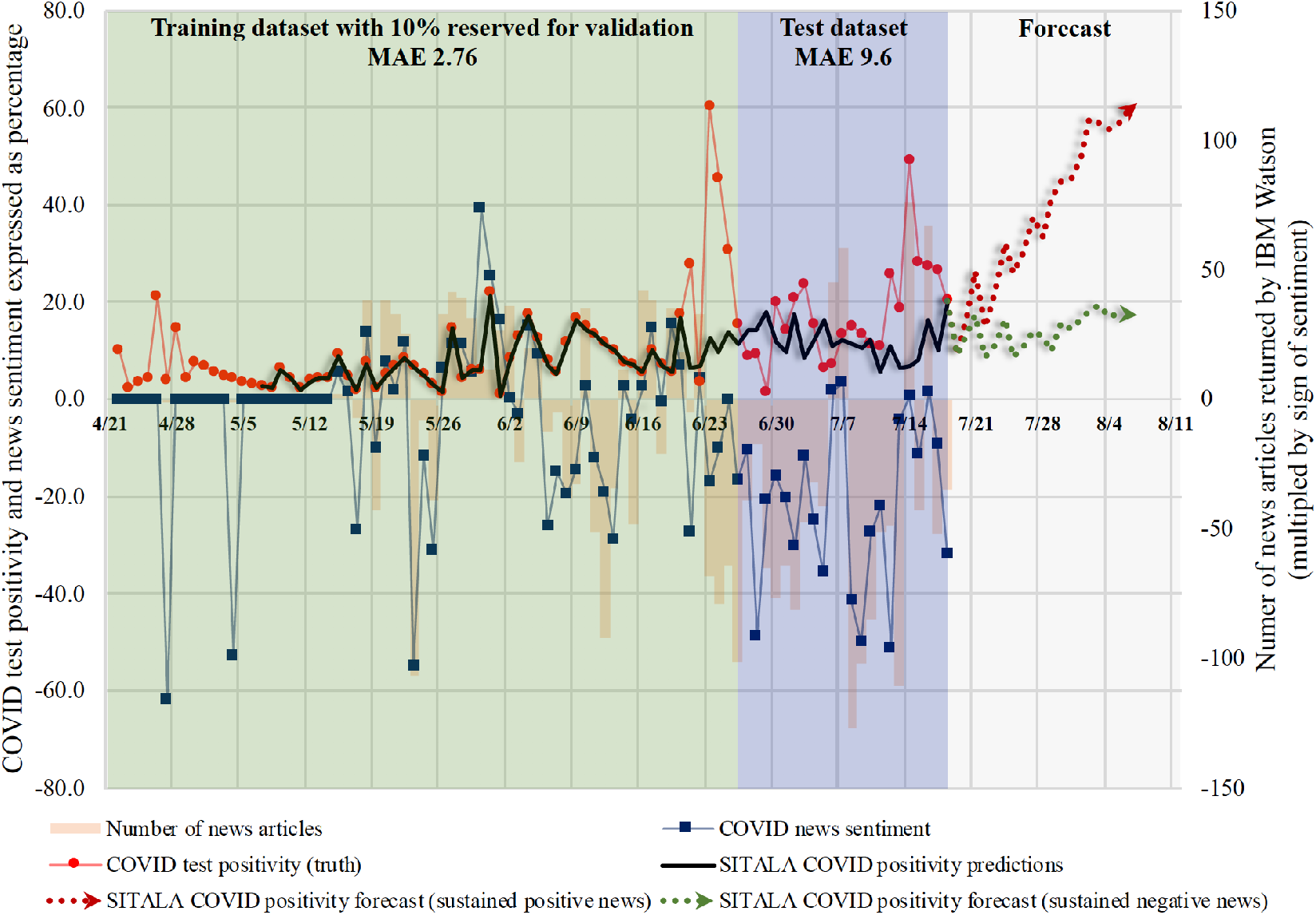
SITALA predictions and relevant COVID-19 data for the Harris county, Texas. Number of news articles returned by IBM Watson discovery, shown on the right axis, started increasing from around mid-May, 2020. COVID-19 test positivity and news sentiment are shown on the left axis. Around 75% of the data (green window) was used for training SITALA, of which 10% was reserved for validation. SITALA was tested on remaining 25% of the data (blue window) for which the mean absolute error (MAE) was 9.6. SITALA forecast (gray window) shows how maintaining a negative sentiment in the news about the spread of COVID-19 can be beneficial to control and eventually decrease test positivity.

The data was divided into training and test datasets comprised of 75% and 25% of the entire data (i.e., 04/21 to 07/17) respectively. 10% of the training data was used for validation. The continuous black line with shadow shows the predictions of trained SITALA over the entire dataset. SITALA is able to capture the response of the COVID-19 positivity data with a mean absolute error (MAE) of 2.76 for training dataset and 9.6 for test dataset. SITALA is unable to capture the highest spikes encountered in both the datasets of COVID-19 positivity. This may have been due to the smaller number of observations in the total dataset (a total of 88 days’ worth of observations is not at the level of the requirements of big data) and smoothing out effect that is introduced by the time window of 16 days.

A maximum positive sentiment of 0.7 and negative of −0.7 were used as the bounds for future news sentiment. With these as the inputs to SITALA, two extreme forecasts were obtained till 08/07. These are also shown in **Fig. 2** with dotted red and dotted green lines with shadow, respectively. SITALA forecasts the positivity of COVID-19 in Houston to lie within this uncertainty cone. A sustained positive sentiment, e.g., “masks are optional”, may prove disastrous for the spread of coronavirus in Houston. On the contrary, a sustained negative sentiment, e.g., “death count for COVID-19 is growing at an alarming rate in Houston”, may help to discourage social gatherings and to keep the COVID-19 positivity under check.

## Discussion

This study highlights the multivariate nature of COVID-19 positivity. The unknowns about the disease have not yet been thoroughly understood. However, public policy makers can benefit from models like SITALA which add the dimension of news sentiment to the positivity data to make forecasts. The long-term effect of sentiment due to the virus incubation period of 14 days can be captured using an AI making use of dilated causal convolutions. News publishers also have a bigger role to play in curbing the spread of coronavirus in Houston. SITALA is a constantly evolving AI and should be enhanced with newer data, as and when available, using transfer learning. SITALA may be deployed at other similar crisis-stricken counties in New York, Florida, and California.

## Limitations

The query searched for the articles having ‘houston’ in the url may have caused omission of few relevant articles that did not have Houston in the url. During the initial few days of training dataset, there were hardly any articles relevant to the IBM Watson query and thus the sentiment during this period was assumed to be neutral, i.e., a value of 0.

## Ethics*(17, 18)*

SITALA may only be used as an ethical guide by public policy makers to set the tone of policies. The author does not support any unethical use of SITALA.

## Data Availability

All data is available in the main text or in the supplementary materials

https://dshs.texas.gov/coronavirus/additionaldata.aspx

## Author contributions

PSD designed the research, performed the research, analyzed the data, wrote and edited the manuscript;

## Data and materials availability

All data is available in the main text or in the supplementary materials.

## Supplementary Materials

### Materials and methods

COVID-19 test positivity data This data is available to the general public and can be downloaded from the website of Texas Department of State Health Service (https://dshs.texas.gov/coronavirus/additionaldata.aspx).

News sentiment data The data for news sentiment was obtained by querying IBM’s Watson Discovery News tool. The tool provides 200 free queries per user per month. More information can be found here: https://www.ibm.com/watson/services/discovery-news/. Discovery News employs natural language processing to return answers to the queries. It also analyzes the sentiment of the news articles. The query used in this study was:

`Is the spread of coronavirus or covid-19 or 2019-nCoV under control?`

Include analysis of your results

`average(enriched_text.sentiment.document.score)`

Filter which documents you query

`publication_date::”2020-05-29”,url:”houston”,(enriched_text.keywords.tex1xt:”coronavirus”|enriched_text.ke ywords.text:”COVID-19”|enriched_text.keywords.text:”2019-nCoV”)`

A sample query along with the output from Watson Discovery News is shown in **Fig. S1**.

**Fig. S1.**
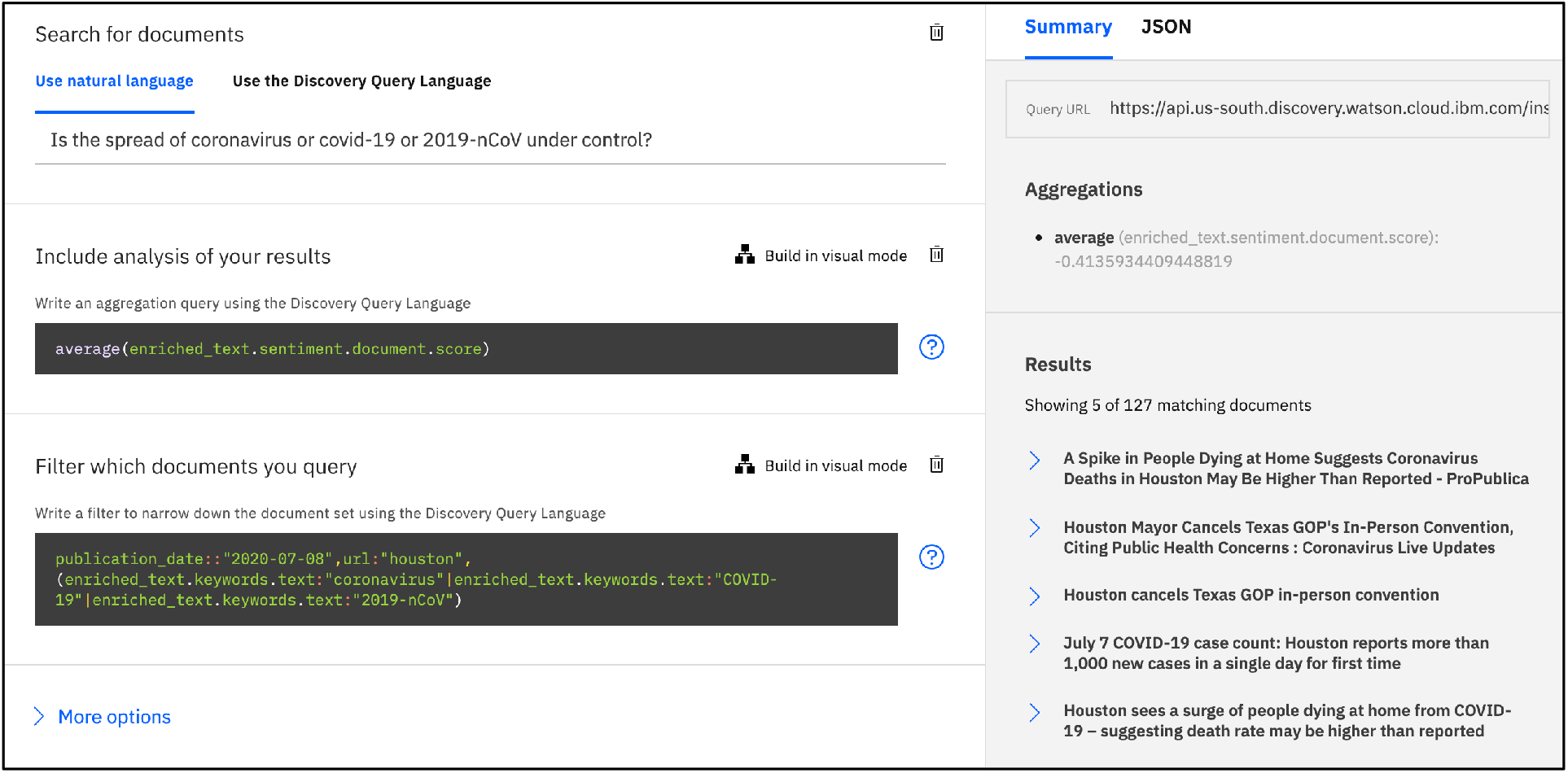
Sample query output from IBM Watson News Discovery. The present study used the exact same query to determine the sentiment of news (-1 implies maximum negative sentiment and +1 implies maximum positive sentiment) over varying publication dates. Location-specific (viz., Houston) articles were filtered using`— url:”houston”`

The entire dataset is reproduced in **Table S1**

**Table. S1.**
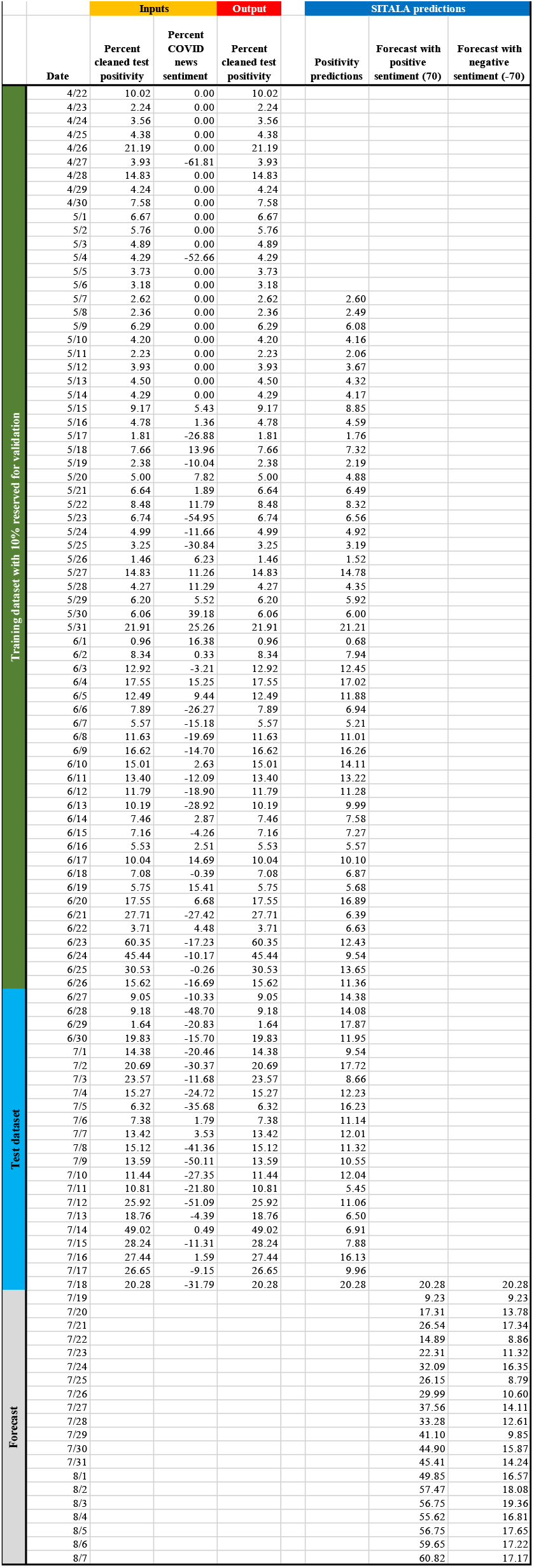
Entire dataset, for the Harris county, that was used in the present study. Also included are the predictions and forecast of SITALA. Window size of 16 was used in this study therefore the predictions of SITALA start from 17^th^ day (i.e., 5/7)

<u>SITALA architecture</u> The python code for the architecture of SITALA is reproduced below. Causal padding and relu activation were used along with 4 dilated convolutional hidden layers. Contact PSD to obtain the trained SITALA for transfer learning.

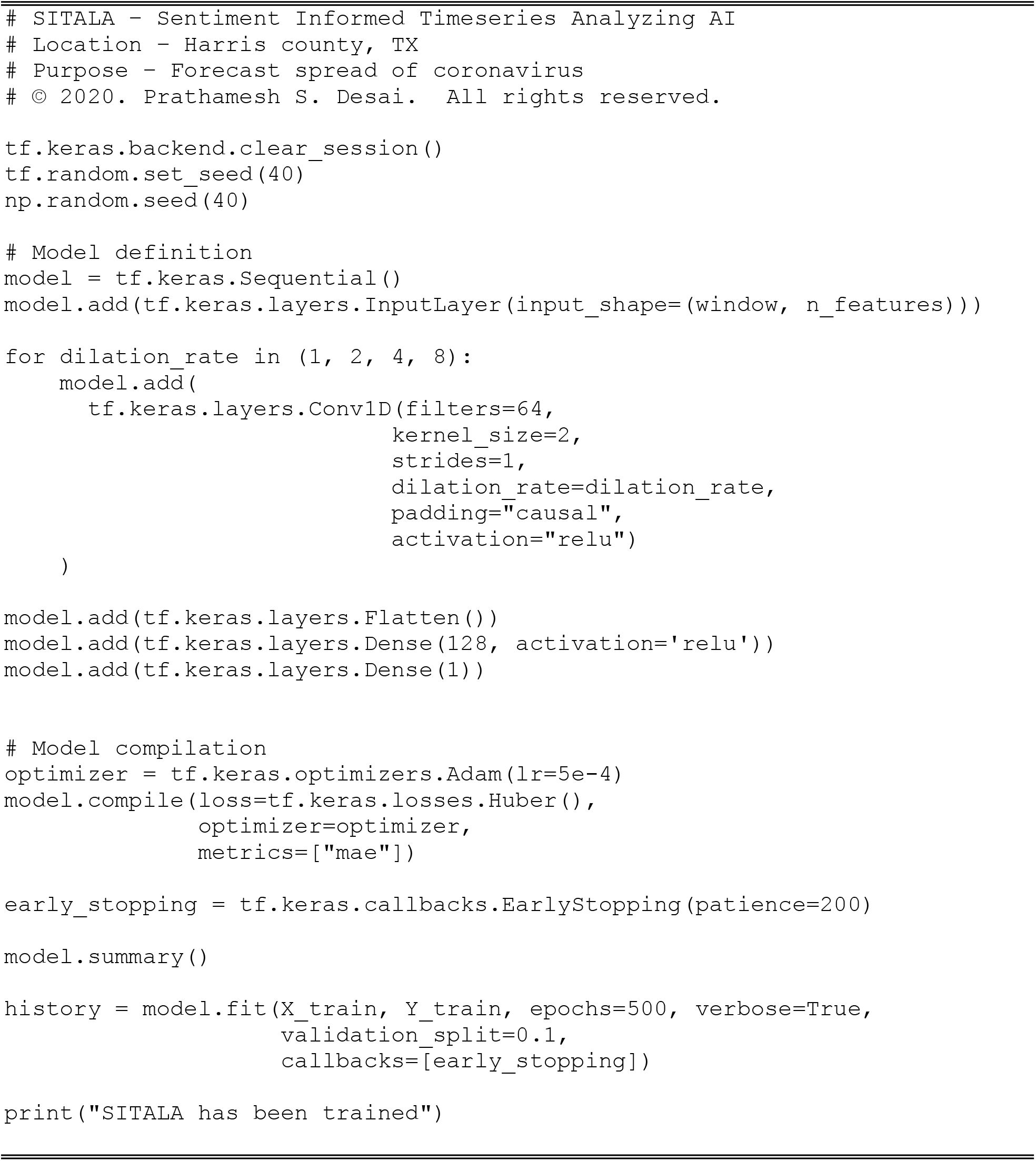

